# A novel comprehensive Tuberculosis (TB) control programme methodology based on the nexus of participatory action research inspired public health and precision treatment approach

**DOI:** 10.1101/2024.01.02.23300347

**Authors:** Shirsajit Mitra, Partha Jyoti Saikia, Anuja Dutta, Rupam Das, Gokul Das, Tutumoni Baishya, Chayanika Das, Tulika Sarma, Sonali Das, Lekhika Pathak, Bikul Das

**Author notes:** These authors contributed equally (Shirsajit Mitra, Partha Jyoti Saikia). Correspondence: Bikul Das, MBBS, PhD. **Abbreviations:** KTC-KaviKrishna Telemedicine Care, Sualkuchi; KKL: KaviKrishna Laboratory; IKS:Indigenous Knowledge System; K-IKS: KaviKrishna Center of Indian Knowledge System IKIN: Indigenous Kamarupa Information Network; CBPR: Community-based participatory research, MDR: Multi-drug resistant, MTB: Mycobacterium tuberculosis.

## Abstract

Tuberculosis is emerging as a major global health issue for the under-developed regions across the world including India’s North East. Importantly, the incidence of multidrug-resistant (MDR) strain of Mycobacterium tuberculosis (MTB), the causative agent of TB is increasing. Furthermore, the disease is associated with stigma, and therefore people’s wholehearted participation in care, and research support is lacking. Here, we report the development of novel research methodologies through a process of participatory action research (PAR) in India’s Northeast region (NER). The region is economically and socially underdeveloped having approximately 10% higher incidences of TB cases reported than the rest of the country. We have taken an indigenous knowledge system (IKS) based PAR approach, where a century-old local philosophy, Vedic altruism (Jiva Upakara Cikitsha Tantra) has been integrated into our research. Our goal has been to integrate philosophy, clinical care, and the latest clinical, microbiology, and cell biology research tools into one cohesive “knowledge emergence” hub, and then connect it with world’s outstanding research hubs to develop a global PAR against TB. Through this unique PAR-based approach, we were able to identify the stem cell niche as an important factor in TB dormancy, reactivation and MDR evolution. Importantly, we also discovered a natural aerosol-based TB vaccination mechanism. In this paper, we describe various methodologies that we have established as a part of the IKS/PAR-based TB research process. These methodologies include the PAR-based clinical care and monitoring, isolation of live MTB from the peripheral blood and bone marrow-derived progenitor cells, culture and drug-sensitivity test, the challenge of MTB DNA extraction and whole genome sequencing (WGS) from a small population of MTB intracellular to stem cells. We have also developed methodologies to detect viable but non culturable (VBNC) MTB of smear-negative pulmonary TB patients, as well as patients suffering from non-tuberculosis mycobacterium (NTM) infections. These methodologies now enable us to detect patients with an early stage of MDR evolution and compensatory mutation by isolating MTB intracellular to circulating progenitor cells. Importantly, the PAR setup now enables us to collaborate with the world’s leading scientific communities to further our knowledge of TB pathogenesis.

## Introduction

Tuberculosis (TB) is a chronic infectious disease caused by the acid-fast bacilli Mycobacterium tuberculosis (MTB) that poses a major global health threat (1). Although global TB cases have reduced in the past years, more than 10 million people are estimated to have active TB as in 2022, according to the WHO’s latest report (2). After the Covid-19 pandemic, TB related death was temporarily reduced due to the lack of reporting, but the incidence and morbidity associated with TB is increasing in the post-Covid19 reporting, becoming higher than AIDS in the global infectious disease trend (2). The ubiquitous and dormant nature of this disease’s pathogenesis makes it difficult to completely eradicate the disease (3). It is estimated that more than one-third of the global population has dormant MTB (dMTB), and that 10% of these cases pose a risk of developing into clinically active TB cases during the person’s lifetime (4). In the last eight decades, several control programs to eradicate TB have been implemented, and yet TB has remained a significant public health issue in developing countries such as India (5). Hence, keeping in line with the UN’s vision of eliminating the TB epidemic by 2030, it is of utmost priority to now develop a novel and effective TB control program.

The TB control programme comprises several steps: patient screening, diagnosis, treatment, follow-up, and continued evaluation for detecting potential MDR TB relapse cases (5). One of the primary reasons for the unsatisfactory outcomes of TB control programs in India and other nations is their generalised approach (6). Therefore, the patient recovery rate in lieu of the TB control programme treatment protocol varies from region to region. Major concerns in these control programs lie in a lack of effective monitoring over various factors such as the identification of mechanism and site of MDR-TB evolution and its management, dormant TB reactivation, the diverse host-pathogen interaction, and circulating MTB lineages. For example, several comparative scientific studies have reported the different strains circulating in different parts of India (5)(6), and yet, the TB control program has so far failed to address unique characteristics of these strains, their dormancy and mechanisms of drug-resistant as well as spread in the community. In addition to these limitations, TB control programs encounters periodic global health threat such as HIV/AIDS, and the recent outbreak of Covid-19. According to the WHO’s latest report on tuberculosis, which states the number of global TB cases have increased since Covid-19, the Covid-19 pandemic has been detrimental for the TB control program, (2). Additionally, studies have reported that Covid-19 infection can enhance the chances of TB reactivation, which will lead to more TB incidences globally in the near future (7)(8)(9). In addition to these limitations, the control programs lack feedback from the community-based research group, and so far remained a top-down, centrally controlled approach which limits the active participation by the community.

To address the above limitations, KaviKrishna has been taking a participatory action research (PAR) based approach for the last three decades. Our goal has been to develop a novel approach of TB patient screening, treatment, and disease progression/dormancy monitoring utilizing an Indigenous Knowledge System (IKS) based public health approach method along with modern molecular and cellular biology methods including the recently introduced sequencing analysis and stem cell evaluations. Thus, we have taken a participatory action research (PAR) approach to develop a community-based participatory research (CBPR) project to manage TB patients in a rural area of the Kamrup district in Assam, India (10). The IKS/PAR includes patient monitoring and evaluation, providing anti-inflammatory nutrition supplements, such as un-denatured whey proteins to poor patients suffering from TB associated malnutrition.

PAR is an effective method to integrate IKS for the new knowledge generation beneficial for the community as well as the growth of basic science in a specialized field of research (11)(12)(13)(14). PAR methodologies have also been used for social networking analysis by us and others (15)(16) Using this social network analysis-based CBPR approach, a pilot study was conducted primarily in the temple complex of the rural Sualkuchi-Hajo region, in Kamrup district, Assam, India. It has a total population of 50000 and a very high population density of 3000/km^2^. Notably, we have incorporated an indigenous philosophy of Vedic altruism, the Jiva Upakara Tantra in our PAR methodological process (17), and as per interaction and recommendations by the indigenous philosophers, we have developed not only a clinic, but also a sophisticated microbiology and stem cell laboratory in a rented facility of nearby high-tech research institute, the Indian Institute of Technology-Guwahati. Furthermore, as per PAR’s evolving networking methodologies, we have developed collaborative projects with leading national and international research institutes including Stanford University, USA, Harvard University, USA, Jawaharlal Nehru University, New Delhi, India, Gauhati University, Guwahati, India, and Indian Institute of Technology-Guwahati, India. Importantly, to facilitate the network we have expanded our PAR based research into the New England communities through our sister lab, the Thoreau Lab for Global Health at M2D2, University of Massachusetts, Lowell, USA.

In this methodology paper, we now describe various methods that had been developed through our PAR approach at the KaviKrishna through its several independent units, the KaviKrishna Telemedicine Care (KTC), KaviKrishna Lab (KKL), and the KaviKrishna Centre of Indian Knowledge System (K-IKS). Our work at K-IKS and KTC has identified a social network system, the Indigenous Kamarupian Information Network (IKIN) prevalent among the indigenous communities of Sualkuchi-Hajo cultural complex of Kamarupa, Assam, India (15)(17). We then incorporated IKIN as a social networking method where people can communicate on an open platform and exchange traditional knowledge among them leading to knowledge emergence (10). Utilising the IKS-based CBPR approach, we previously conducted a TB dormancy study amongst the rural indigenous Idu-Misimi tribe of Arunachal Pradesh and reported that dormant MTB (dMTB) reside inside BM-MSCs of successfully treated TB patients (18). Our team also completed a contact TB investigation study of 15 years in a rural population of Kamrupa to understand the transmission strategy of smear-negative TB subjects (19). In this manuscript, we will thoroughly describe the methodologies utilized for these studies, and associated pre-clinical studies.

Yet, another concern is the potential site of MDR evolution in post Anti-tubercular treatment (ATT) TB cases and its transmission basis in the given community(20)(21). Several studies have reported that resistance to a first line anti-TB drug Rifampicin (RIF) in MDR-TB strains and is the most common clinical finding worldwide (22)(23)). Mutations in the RNA polymerase beta-subunit (rpoB) gene provide RIF resistance in MTB (24). The mechanism of MDR-TB evolution may result from rpoB mutations and/or inherent resistance to other drug resistant genes (isoniazid), such as drug modification by MTB enzymes and the presence of a drug efflux pump (20)(25)). Moreover, mutations in the rpoB gene causes fitness loss, which the pathogen compensates for by secondary mutation in the rpoA and rpoC genes, known as compensatory mutation, which cause fitness gain (26)(27). The rpoC mutation is the most prevalent compensatory mutation (26)(27). The epistatic interaction between rpoB and rpoC may result in MDR-TB development and transmission in the community (20). Currently, there is a dearth of studies elucidating the basis and site of the evolution of non MDR to MDR-TB in relapse TB cases. The matter is made more complicated by our clinical observations that sputum-negative pulmonary TB subjects may later on exhibit MDR evolution. We have developed in vitro assays to study the viable but not culturable (VBNC) MTB present in the sputum of subjects with sputum-negative PTB (manuscript under preparation). In this subjects having VBNC in the sputum, we recovered viable MTB from BM-MSCs and circulatory CD271+ progenitor cells. Importantly, our preliminary data reveals that MTB recovered from BM-MSCs and circulatory MSCs of subjects with sputum-negative post-anti-TB drug-treated relapsed cases have mutations in the rpoB and rpoC genes. These results imply that the stem cell niche may play a role in MDR evolution (manuscript under preparation). As per our preliminary findings, we speculate that the hypoxic BM stem cell niche can be a putative site to evaluate for early detection of MDR evolution (21). Furthermore, gauging the early MDR evolution within a selected patient cohort of a certain area or community will provide insight into the lineages of circulating MDR-MTB strains and their viability rate. Based on our findings, we postulate that this TB control program approach will have a positive outcome based on its successful pan-India implementation.

Thus, the PAR based setup of this clinical and laboratory research facility uniquely positions us to collaborate with the world’s scientific communities to develop an IKS-based TB control program that can develop a precision treatment approach. Significantly, through nearly one and a half decades of non-profit and CBPR-based investment in research, we have been able to achieve the technical expertise in studying the newly discovered role of stem cell niche defense in MTB dormancy and reactivation.

In this manuscript, we will thoroughly describe the methodologies utilized for the above studies including our maturing capacity building of bed-to-bench site research including the detection of VBNC in the sputum negative samples, and doing whole genome sequencing (WGS) and bio-informatics pipeline, plus depositing the WGS data in the international repositories. We strongly believe that these capacity building achieved in a remote but TB-endemic area of India will help our global network to work with us for the evolution of a novel, PAR-based TB eradication platform as envisioned by our philosopher elders of Vedic altruism (17)(19).

## Methods and Outcomes

### 1. Ethical Approval for Clinical Study

The proposed protocols for clinical study were approved by KaviKrishna Clinical Research Review Committee and KaviKrishna Institutional Ethics Committee. Necessary consents from the selected patient subjects of homogenous Sualkuchi-Hajo area population were taken prior to induction for the clinical study. DOTS II individuals were recruited prospectively based on data from the local TB control programme and chest X-ray findings. Following informed consent, 6 to 7 ml of BM was collected from the posterior superior iliac crest, and the immune-magnetically sorted cells were lysed and subjected to DNA isolation using uMACS technology, as described below. Additional consent was taken for publishing the clinical research findings.

### 2. Developing an IKS-based telemedicine network in rural Kamrup for TB patient screening, monitoring and conducting clinical research

For better control of TB spread in a community, participatory action research (PAR) has been tried with varying results (28)(29). However, so far, the IKS based PAR has not been tried to develop novel methodologies for knowledge generation about the control and eradication of TB. Northeast India has many diverse groups of indigenous people living in rural areas (30). As such, a robust rural public health approach is crucial to serving these populations and thus, is a critical element of rural health development. In India, the general health of the people in rural populations is inferior to that of urban populations (31); rural areas tend to have shorter life expectancies and high mortality rates due to infectious diseases like tuberculosis (TB)(32). Despite these adverse statistics, while 72.2% of Indian people reside in rural areas, only 40.8% of healthcare workers are available for rural areas (33). Thus, it is imperative that public health management of rural populations in developing countries improves.. Accordingly, we planned to develop an IKS-based telemedicine approach in the rural Kamrup area to conduct a population based TB reactivation study.

An Indigenous Knowledge System (IKS) is an adaptable knowledge system created and maintained by an indigenous culture over many generations. Our lab (KKL) has been working with a rural population from the Sualkuchi-Hajo area of Assam via KaviKrishna Telemedicine Care for the last 25 years (18)(19). We are inspired by a historical communication system used by the local silk weaving community of Sualkuchi, as well as the metaphysical properties of their traditional philosophical system, the Vedic Jiva Upakara tantra (15)(19). The core components of their communication system are: the setup of a skill-development hub for skill practitioners, and the establishment of a communication network through local temples, known as the indigenous Kamarupa information network (IKIN) (15)(18). As shown in the Figure 1, the IKIN works through the connection between indigenous temple complexes, which are traditional meeting place for communities.

**Figure 1:**
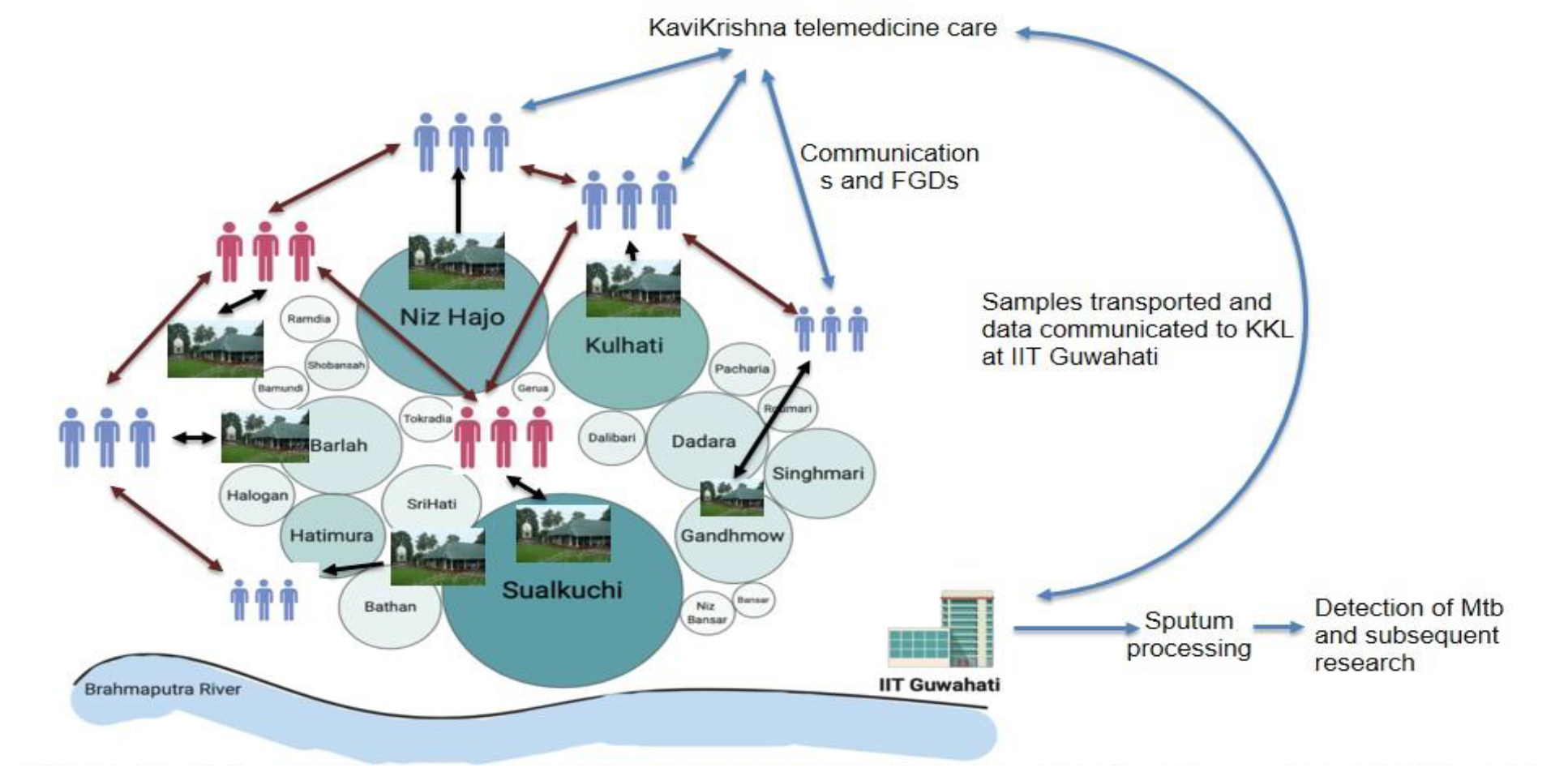
Depiction of IKIN based network in greater Sualkuchi Hajo area with students community being active and in constant to and fro communication with satra temples active in the area (communication with satra and students are depicted with black arrow heads) for detection of active TB cases among regular temple visitors and other in the nearby society. Talks and FGDs are held at KTC with the students to gather information from the different satra temples (maroon colour arrow heads depicts communication among active student group in different areas and blue colour arrow heads depicts to and fro information sharing among student groups and KTC). On the basis of students information, samples are collected by home visit and are transported to KKL AT IIT Guwahati for subsequent clinical test and scientific, academic research.

Sualkuchi has 25 Kirtan temples with a closed door set-up called “Namghar.” In the Hatisatra Namghar, established in 1650 AD, 40-70 devotees between the ages of 40-80, both male and female, gather twice daily for the community-chanting of Kirtan. Many devotees attending this Namghar seek medical treatment from KaviKrishna Telemedicine Care (KTC) (19). KTC has been screening TB patients among these devotees using this temple network since 1994. While managing PTB subjects in rural India through the KaviKrishna Telemedicine Care (KTC), we have simultaneously discovered the strength of Vedic altruism based philosophy of medicine (10,19), and its possible application in rural tele-health, a rapidly emerging field in rural health (34). Notably, we have uncovered an IKS-based aerosol-inoculation practice reported by the British colonial administration (19) that demonstrates the possibility of the emergence of aerosol inoculation based herd immunity against tuberculosis (TB), where patients with pulmonary TB (PTB) were asked to perform kirtan chanting in the Namghars to transmit good-nigudahs around the community. We have utilized this IKIN network to trace the TB patients and their contacts with TB symptoms. We have also developed IKIN based digital health device comprised of Nigudah yoga (36), focused group discussions (FGDs), providing whey protein supplements, and regular health monitoring (37). Using this IKIN network, we developed a TB control protocol, and in the process of extending our activities to the entire Northeast through the numerous temple networks, as well as community non-profits such as RIWATCH of Roing, Arunachal Pradesh, India. Since 2010, we screened 300 PTB cases/500 suspect cases from these temple complexes, after recruiting student volunteers from local colleges. These student volunteers traced the patients residing in their locality, and also their contacts by taking the help of the local temple network. Thus, a Community Based Participatory Research (CBPR) approach was started, where the stakeholders included the local temple network, the student volunteers, our doctors, our telemedicine clinic members, lab research students and most importantly, the patients and their families. At first, the volunteers were trained regarding general awareness about TB, its spread, control and preventive measures, dangers of MDR TB. The temple network was also involved in encouraging and mobilising the patients and their caregivers to participate in this awareness drive. Additionally student volunteers were taught the practical aspects of TB diagnosis, treatment and control like collection of sputum, proper disposal of biomedical waste, donning and doffing of PPEs while dealing with sputum smear patients etc. After a post training assessment, volunteers were stationed in various locations across the Sualkuchi-Hajo area, to help spread the awareness of the TB disease and ways to prevent spread by distributing pamphlets, performing home visits, and conducting focused group discussions (FGDs). After obtaining informed consent, all study subjects were screened and identified by observing the clinical symptoms for TB, as well as sputum-examination reports obtained from the local hospital as described in a study (19). The study was implemented by field-based contact investigation including home visits, temple visit and then will be confirmed with their concerned physicians and health workers (manuscript under preparation). The details of the study subjects were maintained in the lab register using a unique identity number and followed up with regular intervals. Utilizing the same temple network, the subjects are under the monitor of KTC for probable relapse or MDR evolution. (38). An example of the data set of our patient follow-up is given as a supplementary file, Mitra et al. TB patient follow up KaviKrishna telemedicine care. In this data set, we provide information of 210 patients.

### 3. TB patients’ sputum, bone marrow and blood collection from the rural population for a longitudinal study

We have been doing the sputum collection and AFB staining since 1994 (19), and also took this experience to Bhutan’s TB control program at Mongar Hospital during 1995-1998 (38), where the first observation of TB hiding in bone marrow was observed (38). In 2010, we recruited subjects from the Idu-Mishmi tribe living in a remote mountain range of Arunachal Pradesh, India utilising our KTC network for a TB dormancy study (18)(35). All participants were older than 40 years and gave written informed consent. Participants were recruited by RIWATCH with BD’s direct supervision. Previously treated subjects were prospectively recruited based on the local tuberculosis control program data and Chest X-ray findings, as described (37). Subject who were pregnant or had diabetes or heart disease, were excluded. All participants had routine HIV testing using Abbott Determine HIV1/2 rapid antibody assay test kit (Abbott Laboratories). The Idu-Mishmi community has so far not been found to be positive for HIV/AIDS. Patients PTB were diagnosed based on sputum and Chest X-ray findings and all subjects had previous history of sputum AFB (Acid Fast Bacillus) +++, and Chest X-ray showing active cavity formation. Patients were given DOTS II by the local government district hospital (managed by IP and patient information was studied by BD and DK). After completion of treatment, AFB and culture of sputum were performed by the Ranbaxy Laboratory, Mumbai and were found to be both negative. In addition, no MTB-DNA could be detected in treated patients’ sputum, blood and BM aspirates. Healthy control participants were recruited from volunteers of 40-50 years old. After proper consent, 6-7 ml of bone marrow was taken from the posterior superior iliac crest following local anaesthesia as per standard procedure with proper aseptic conditions. The marrow was diluted in PBS containing 5% FCS and BM-MSC was obtained as described (18). We obtained 2.3±0.7 x107 BM-MSC per ml of BM aspirate which is consistent with previous reports (18).

Now, we are following up these patients for dormant TB reactivation and detection of circulatory MSC containing intracellular MTB. Since 2018, the KTC has increased the use of digital tools for our patients, and volunteers to help patients. We have been following up these patients with the help of KTC volunteers. Recently, following a DBT funded grant, we recruited n=250 TB patients, including MDR-TB, from the entire northeast region for another study to evaluate the stem cell’s role in MDR evolution. In both of these populations, we are currently applying the Vedic Altruism based telemedicine approach for continuous monitoring for probable relapse. We are now using a whole genome sequencing (WGS) based surveillance approach to monitor the evolution of MDR TB and evaluate the role of the stem cell niche. The samples including sputum, blood and bone marrow are collected in a sterile container by the trained KTC volunteers and transported to KaviKrishna Laboratory on the day of collection for further process. The transportation was done using a 4⁰C cold maintenance system.

These two longitudinal studies and their success indicate that CBPR based approach in using IKS tool may be an effective method in developing novel TB control programs tailored to the needs of a given community.

### 4. Sputum sample processing, identification of MTB, culture and DNA extraction

In KKL, we have established a sterile enclosure and installed a Biosafety Cabinet-3(Figure 2A) as part of the DBT grant objective. Standard BSL-3 grade SOPs are strictly followed at all-times for this study. We processed the sputum samples using the NALC NAOH approach, based on multiple conventional procedures for sputum decontamination. First, each sputum sample was vortexed for 5 minutes with an equal amount of NALC NAOH solution. After 15 minutes, phosphate buffered saline was added and the volume in a falcon tube was made up to 50 ml. After adding PBS, the samples were centrifuged at 3000 RCF for 20 minutes at 4⁰C. The supernatant was discarded, and 1.5 ml of 7H9 media supplemented with 10% glycerol was added to the pellet (7H9 media supplemented with 10% glycerol is utilised as a storing medium for decontaminated sputum samples). In duplicate, 100-200 microliters of storage media were inoculated in Lowenstein Jenson medium slants and incubated at 37°C.The slants were monitored for 8 weeks to see if colonies grew in the cultures (Figure 2B). Once colonies showed luxuriant growth in the slants, they were selected for Ziehl-Neelson staining and examined under a microscope for the presence of acid fast bacilli (AFBs) (Figure 2C). After confirming the presence of AFBs under the microscope, colonies were scooped for DNA extraction. Initially, we opted for the standard company provided DNA isolation kits. However, the quality of DNA obtained with the kits was not ideal for WGS. Also, keeping in mind the cost-effective aspect of rural healthcare, instead of using the expensive MTB DNA Extraction kit, we proceeded with an optimised version of the traditional phenol chloroform extraction procedure for MTB DNA isolation from clinical isolates (manuscript under preparation). The isolated DNA samples were kept at - 20°C until further usage. For DNA quality check, Agarose gel electrophoresis and Nanodrop spectrophotometer reading approach was followed.

**Figure 2:**
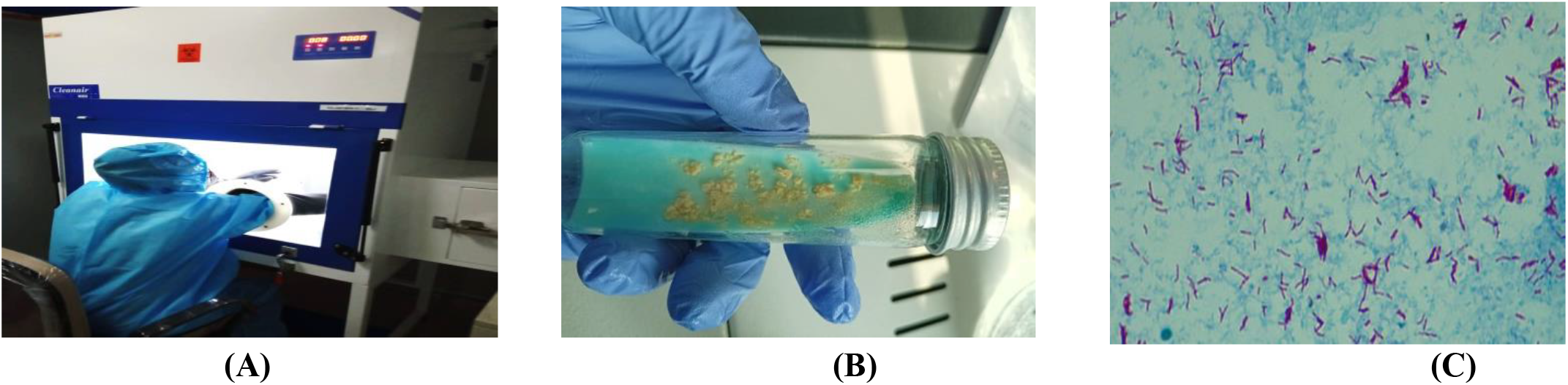
Images depict A) Sputum sample processing and MTB culture carried out in Bio Safety Cabinet 3 at KKL; B) MTB colony growth observed in LJ slant; and C) AFB slide observation under microscope.

## 5. Drug susceptibility tests using proportion method

### We have established the proportion method of DST as described by WHO

(39), and as per the SOP given in the WHO website (40). This method was first described by Canetti et al (41). Briefly, MTB grown in LJ medium containing a concentration series of drugs such as rifampicin were observed for growth inhibition. The percentage of growth (number of colonies) of a defined inoculum on a drug-free control LJ medium was compared with the LJ containing the critical condition of an anti-TB drug. Two methods are used, the indirect (using a pure culture of MTB that yield >50 colonies; freshly sub-cultured H37Rv serves as control strain), and the direct method (decontaminant sputum grading of 2+ or 3+). We have established the indirect method of DST as follows.

### Preparation of LJ slants with Rifampicin

The LJ slants were made using rifampicin at a concentration of 20 ug/ml, as opposed to typical LJ slants. Isoniazid dose 0.2 ugm/ml, ethambutol 2 ug/ml and 4 ug/ml dihydrostreptomycin.

### Preparation of inoculum

Using a loop instrument, a tiny representative sample of about 4-5 mg was removed from the primary culture. This sample was then placed in a McCartney bottle containing 1 ml of sterile distilled water and six 3 mm glass beads. A vortex was used to vigorously mix the bottle for roughly 20-30 seconds. An extra 4-5 ml of distilled water was gently added to this mixture while the bottle was continuously shaken. Larger particles are allowed to settle. The next step was to carefully pour the mycobacteria into a new sterile McCartney bottle, clear of these gritty particles.

The bacterial suspension’s density was modified by gradually adding distilled water until it reaches a concentration of 1 mg/ml of tubercle bacilli, determined by comparing it with the McFarland standard No.1.Suspension preparation for the economic variation of the percentage method: 1 ml SDW with six 3 mm glass beads + 1 loop full (3 mm internal diameter) of culture vortexed for 20 - 30 seconds is neat. 4 mL of SDW was added to the aforementioned solution. SDW adjusted turbidity with McFarland 0.05. S2-10 −2 2 tidy loopfuls + 2ml SDW S4-10-4 2 loops of S2 + 2ml SDW

Shaking was used to combine the components. For rifampicin medicines, two slopes of medium without drug and one slope of media with drug were inoculated with a loopful of each dilution.

### Preparation of McFarland NEPHELOMETER BARIUM SULFATE STANDARD No.1 (Paik, G. 1980)

By dissolving 100 mg of anhydrous barium chloride in 10 ml of sterile distilled water (SDW), a 1% aqueous solution of barium chloride was made. Similarly, made a 1% solution of sulfuric acid (AR) by adding 0.1 mL of AR to 10 mL of SDW. The 1% barium chloride solution was then combined with 9.9 mL of the 1% sulfuric acid solution. This combination produced the McFarland standard No.1, which corresponds to a concentration of M. tuberculosis of 1 mg/ml. Finally, the tube was parafilm-sealed and prominently identified as McFarland standard No.1, along with the date of preparation.

Incubation and data interpretation: The LJ slants were incubated at 37^0^C, and examined for contamination after 1 week of incubation. If not contaminated, the LJ slants were subjected to data reading after 4 and 6 weeks of culture. The growth of 50-100 colonies on the drug-free media was considered as +, 100-200 colonies as ++, and 200-500 colonies as +++. Every new batch of the drug-containing LJ medium was subjected for quality control by using the H37Rv strain.

## 6. PCR of clinical isolates and culture-isolated DNA sample to rule out Mycobacterial Cross-reactivity and MDR-TB evaluation

Several NTMs mimic similar symptoms as pulmonary tuberculosis.. Moreover Zeihl Neelson staining doesn’t help in easy differentiation of MTB from NTMs. Therefore, it is difficult to distinguish between the two, especially while treating sputum/smear negative cases. Previous epidemiological studies have reported cases of misdiagnosis of NTM led pulmonary infections with pulmonary Tb (41)(42). Among the 170 different NTM species, we mainly focused on the two NTM species *M.abcessus* and *M.avium complex.* These two are primarily known for NTM led pulmonary infections. Interestingly, we found NTM positive samples which were initially suspected to be pulmonary TB cases.

Hence, to rule out the chances of non-tuberculosis mycobacteria and cross reactivity, all decontaminated samples and isolated DNA samples were subjected to PCR. PCR was done with primers specific for *Mycobacteria spp, M.abcessus* and *M.avium complex*, which were commonly reported previously (43)(44)(45). Eventually, we also found evidences of NTMs from patient samples that were initially diagnosed with PTB. Interestingly, our lab also experimentally confirmed the presence of NTMs such as *M.avium complex* and *M.abcessus* through PCR and WGS (manuscript under preparation) Furthermore, PCR amplification of *Mycobacterium spp*. culture isolates and reference strains was done using the primer set HSP N3 and HSP N4, amplifying a 300 bp area of the hsp65 gene (46). A second primer set, ABC T1 (5′-GCAGCATTGAGGTACTTGGAC-3′) and ABC T2 (5′-TCGGTGAGACCCAAGGTGTC-3′), was used to amplify a 190 bp area of the gene Rv1458c. Rv1458c is a gene that encodes a protein that is thought to be an ATP binding ABC transporter (47).The presence of MTB in the cultures was confirmed by amplification of both primers (Figure 3).

**Figure 3:**
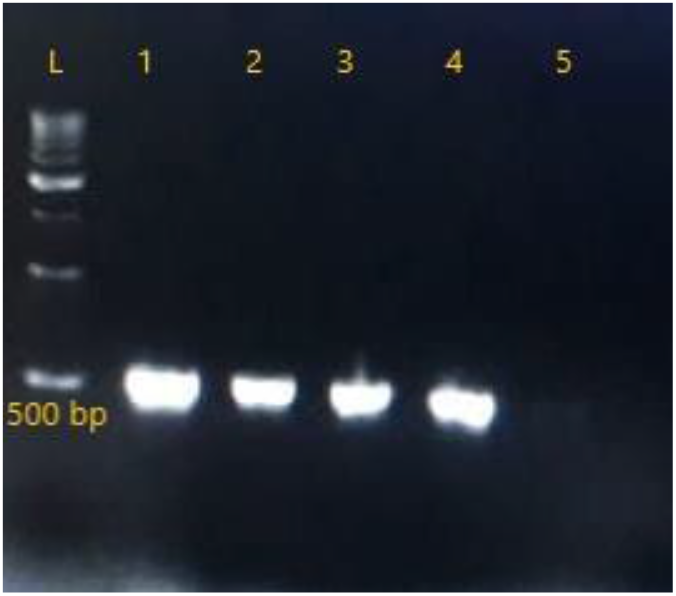
MTBC confirmation using 2% Agarose Gel Electrophoresis post-PCR: L-ladder; 1:Positive control; 2:Patient Sample ID: KKL-15; 3:Patient Sample ID:KKL-24; 4:Patient sample ID: KKL-35; 5:Negative control.

In the follow-up step, PCR +ve MTB samples were further subjected for the identification of MDR/non-MDR strains. MASPCR was used using two sets of primers specific for mutated rifampicin resistant genes: rpoB516, rpoB526 and rpoB531 as described (48).

## 7. Studying the stem cell basis of TB dormancy and reactivation from clinical isolates and in-vivo models

Earlier, we hypothesized that the BM stem cell niche harbouring MSCs and HSCs protects dormant MTB from anti-TB drug therapy by utilizing the innate stem cell niche defense mechanism. Furthermore, we identified that during oxidative stress, MSCs and HSCs exhibit a “enhanced stemness” (state of self-renewal and self-sufficiency) phenotype including the activation of the HIF-2 stemness pathway and a transitory decrease in p53 (49). Fitness gains result in the reprogramming of MSCs and HSCs, which are referred to as ASCs. In the milieu of hypoxia/oxidative stress, these ASCs can exhibit niche modulatory features and serve as an ASC niche defence mechanism (50). Recently, we reported that MTB pathogen may exploit this stem cell niche defense mechanism (21) to enhance their fitness in the hostile microenvironment of lung for PPTBL development or TB reactivation. The dMTB harboring BM-MSCs may migrate to lung following tissue injury or inflammation utilizing its circulatory characteristics. Hence, elucidating the host-pathogen interaction in both dMTB reactivation and MDR evolution is of great importance.

To conduct this study, sorting of the MSC and HSC subpopulation is essential. Hence, keeping in mind the pan-India scenario and potential logistical and handling issues, we used an unique immunomagnetic sorting method instead of the flow cytometry based sorting. The MSCs(CD281+/CD133+) and HSCs(CD133+/CD34+) were first isolated from the PBMCs of human BM and blood samples and were cultured in serum free media, followed by the phenotypic characterization of CD281+/CD45-sub-population as described earlier (18). The sorted CD281+ sub-population cells were then subjected to isolation of mammalian RNA and bacterial DNA for evaluation of residual dMTBs using the uMACS technology as described (18). The presence of MTB DNA was investigated by qPCR using MTB specific primers (18). As a negative control, DNA was obtained from CD281+ BM-MSCs and CD281−/CD45− cells of healthy volunteers from non-endemic areas. The isolated cDNA samples obtained from BM cells were then subjected to QPCR using TaqMan Gene expression assays as described (18) for analysis of the stem cell related gene expression.

Hence, to elucidate the MTB dormancy-reactivation potential, long-term follow up of a particular patient cohort is essential. For this, we utilised the previously established IKS network (15)(19), and applied the CBPR approach for patient selection, sample collection and patient evaluation at regular intervals.

Furthermore, several studies previously evaluated the residing dormant(latent) MTB and its reactivation potential using different *in-vivo* models (51)(52)(53)(54). Hence, to validate our hypothesis of viable dMTBs residing in the hypoxic BM stem cell niche, we concomitantly with the studies comprising clinical isolates and *in-vitro* subjects, designed an *in-vivo* mouse model of non-replicating MTB infection (18)(55). The model was designed using a streptomycin dependent auxotrophic mutant of MTB (strain m18b). The model was used to evaluate the viability retention capacity in *in-vivo* MSCs as described (18)(55). Interestingly, using this mouse model, we were able to demonstrate that dormant MTB could be isolated from MSC populations present in both the lungs and the BM of sick mice which were devoid of streptomycin treatment for 6 months. Viable MTB was obtained directly from CD281+ BM-MSCs in BM. We also used GFP-labelled MTB to confirm infection of this cell type. In the lung, viable MTB was derived from the MSC-enriched SP cell population. When these infected MSCs were re-injected into healthy mice, the animals developed lung granulomas, indicating that the non-replicating MTB preserved intracellularly in the MSCs had the ability to re-infect.

Moreover, as per our hypothesis of reprogramming of MSCs to ASCs gaining an innate stem cell based niche defense mechanism during stressed condition, we tested the reactivation potential of dormant MTB using this m18b mouse model. Parental coronavirus strain MHV-1 was infected in the m18b mouse as described (7). Post co-infection with MHV-1, Viral colony forming unit (CFU) and MTB-CFU assay was performed as explained (7). We found that, on viral infection it led to oxidative stress resulting in a state of enhanced stemness reprogramming. The dMTB harbouring circulatory MSCs respond to the stress microenvironment in the lung by altering itself into ASCs leading to a transient phenomenon of high HIF-2a and suppression of p53. Now these dMTB harbouring ASCs (previously MSCs) while mediating the innate defense mechanism released the residing MTB (now active) (7). The ASC reprogramming was confirmed based on the expression of the related stemness genes Oct-4, GCL, ABCG2 (49) using real –time quantitative PCR assay as previously explained (7). Furthermore, the active MTB was found to successfully infect in the pulmonary site forming Tb granulomas. These findings imply that activated dMTB released by reprogrammed MSCs invaded into the non-MSC lung cells, causing TB disease reactivation.

## 8. Identification of prevailing MTB MDR/non-MDR lineages by MIRU-VNTR and spoligotyping, corresponding drug resistance and compensatory mutation profiles by WGS

Amongst the seven lineages of the Mycobacterium tuberculosis complex, the presence of each of them varies from region to region of India. Using the MIRU-VNTR and spoligotyping approach, we evaluated the circulating lineages as explained earlier (56). We identified Beijing strain, as well as CAS/CAS1-Delhi and several orphan strains (manuscript under preparation) consistent with previous reports (57). Based on the lineages, the mutation points also vary. Earlier reports also suggest the potential role of certain compensatory mutations associated with reinstating the virulence of the drug-resistant MTB strains (26)(27). As per our studies, we speculate that identifying the putative compensatory mutations corresponding to the fitness gain of the drug resistant genes of individual patients may allow us to elucidate the non-MDR to MDR evolution mechanism (manuscript under preparation). Combining the lineage profile and mutation profile of patient subjects from particular cohorts would allow us to make an epidemiological analysis of the circulating strains and also detect the MDR transmission route in the area. Hence, we proceeded with a pilot study of WGS analysis to identify the specific strains using phylogenetic analysis and the respective mutation profiling. In the pilot study, we wanted to evaluate the feasibility of a CBPR based approach for WGS-based patient monitoring and disease recovery.

The isolated MTB DNA samples were outsourced for Whole genome sequencing (WGS), post QC using Agarose Gel electrophoresis and a qubit fluorometer analysis from KKL. Samples with DNA purity ∼1.8 (260/280) and concentration (1ng/ul) were sequenced in the Illumina platform (Model: NovaSeq 6000) on a 150*2 paired-end read basis. Library preparation was done using the standard Illumina DNA Prep kit. The raw reads (fastq file) generated were then analyzed at KKL for downstream analysis (Figure 5). Reads were scrutinized as per standard quality check (QC) protocol using fastQC (version 0.11.9). Post trimming and read filtering using fastQC, the reads with a phred score>30 were then subjected to further downstream alignment with the reference genome. BWA (v0.7.17-r1188) was used for aligning the reads with the reference genome of Mycobacterium tuberculosis H37Rv (NC_000962.3). Sample reads with alignment percentage greater than 90% were considered for variant calling and variant annotation. Variant calling was done using GATK (version 4.4.0.0) to identify the SNPs and Indels (Figure 5). The vcf files were then annotated using the Annovar tool. The annotated files were then further evaluated for studying the compensatory mutation profile of each sample and subsequent selection of study subjects for the stem cell-based study on stem cell niche defense.

**Figure 4:**
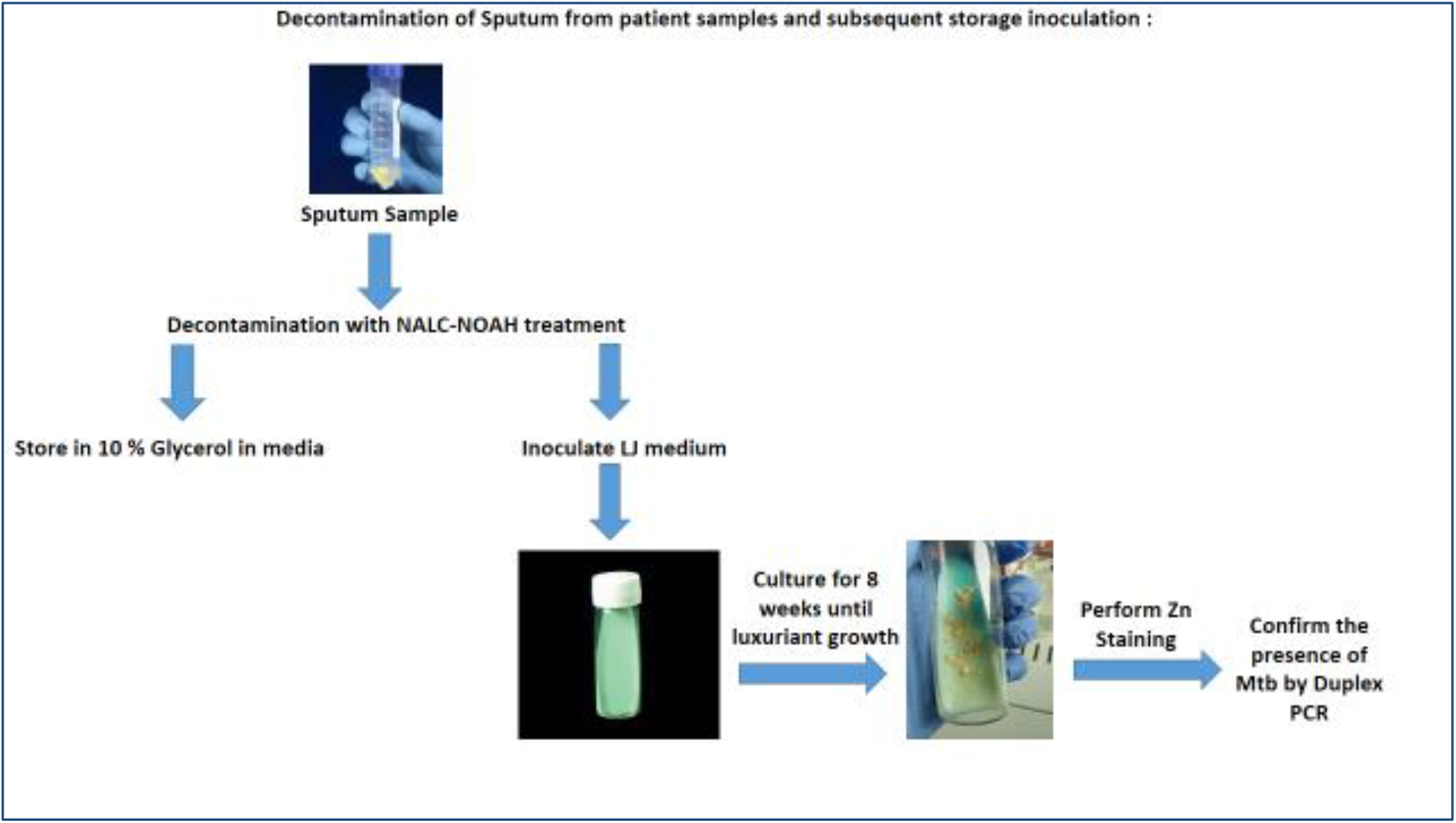
Schematic showing the flow of sputum decontamination to confirmation of culture.

**Figure 5:**
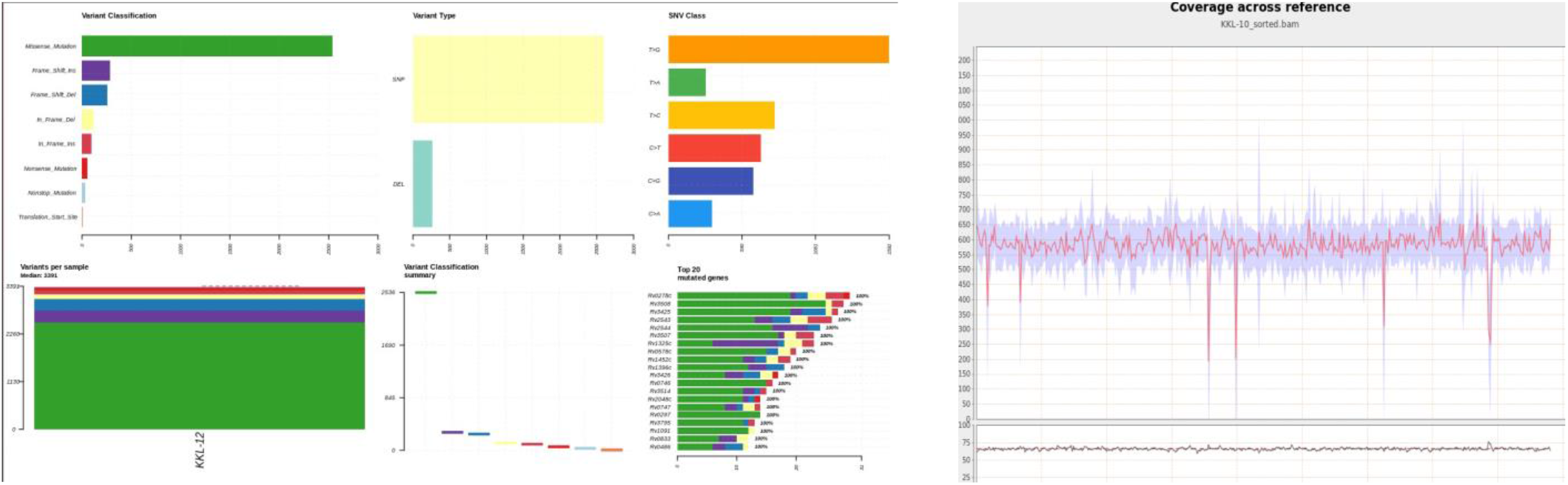
Representative analysis results from the WGS data of one patient sample.

From the post-analysis of the WGS raw data obtained from circulating CD271+ cells, we identified a putative mutation set in the rpoB and rpoC genes of the respective samples (manuscript under preparation). This will allow us to further evaluate the rpoB-rpoC compensatory gain linkage and its role in MDR-TB evolution. The available raw data would be further subjected for MTB phylogenetic analysis using MAFFT tool and IQ-tree tool obtained of each of the samples to identify the circulating strains as described (58). For broader spectrum studies by the whole research community, the raw data of samples will be gradually uploaded in the NCBI online repository. A sample dataset is already uploaded for reference (Accession no. PRJNA1054003).

## 9. Microplate Based Alamar Blue assay (MBAB) method to determine the MICs of drugs and herbal medicinal agents

Minimal inhibitory concentration (MIC) is the in vitro concentration of anti-bacterial agent that a given bacterial strain that shows susceptibility. MIC is the lowest concentration of an antibacterial agent in the in vitro condition expressed in ug/mL. In a strictly controlled in vitro environment, MIC dose can completely prevent visible growth of the tested bacterial strain (63). To obtain credible MIC, proper methodology and competent interpretation of results are required. Two methods are primarily used: dilution and gradient used for MIC estimation. We have used the dilution method (using a liquid broth medium) to obtain the MIC as described (63) using the dye based growth detection method (64). Briefly, H37Ra was prepared in the liquid 7H9GC broth for two to three weeks to obtain sufficient growth. Bacterial inoculum of 5×10^5 CUFs were prepared with the help of 0.5 McFarland suspension (63). Stock solutions of drugs isoniazid (INH), rifampicin(RMP), streptomycin (SM), and ethambutol (EMB) (all obtained from Sigma and dissolved in deionized water except RMP, dissolved in DMSO to prepare stock solutions). Ursolic acid, squalene, and apigenin stock solutions were prepared in DMSO. Stock solutions were diluted in 7H9GC broth twice the maximum desired concentration of sensitivity testing. In a 96-well microplate, 200 ul of sterile deionized water was added to a minimum 25 wells to minimize evaporation of the medium in the test wells during prolonged incubation. Then, the testing wells, as well as control wells received 50 ul of 7H9GC broth. Next, 50 ul of 2x drug or herbal medicine products were added to the testing wells. The control wells received 50 ul of vehicle DMSO of the desired concentration. In these wells, 100 ul H37Ra growing broth containing 5×10^5 bacteria was added. The plates were sealed with parafilm and were incubated at 37C for 6 days. On the day 7^th^, 50 ul of Alamar blue solution (1:1 mixture of 10x Alamar blue with 10% Tween 80) was added to the control wells, and the plates were then re-incubated for 24 hours. If the wells turned pink, the growth was confirmed. Then, the 50 ul Alamar blue solutions were added to the testing wells and then re-incubated for 48 hours. Then, the colors of all the wells were recorded. A blue/violet color was interpreted as no growth and a pink color was interpreted as growth. The MIC was defined as the lowest drug/herbal agent concentration that prevented color change from blue/violet to pink. Note that the MIC results were compared with the Agar dilution assay of Canetti et al using LJ slants. Synergistic interactions between herbal extracts/bioactive compounds and INH + RIF were calculated by estimating the fractional inhibitory concentration (FIC) index. To obtain the index, first, the ratio between the MIC of a RIF+INH was divided by the MIC of INH. Similarly, the ratio of MIC of Ursolic acid + epigenin/MIC of Ursolic acid was obtained. Then, the two rations were added to obtain the FIC. The combination is considered synergistic, additive, and antagonist based on the FIC index as 0.5, 1 or 1.5 respectively.

## 10. Most-Probable-Number (MPN)-Based Minimum Duration of Killing Assay (MDK)

Determining bacterial susceptibility to antibiotics requires not only the MIC estimation but also the estimation of duration and peak of antibiotic exposure required to kill more than 99-99.99% bacterial population in a given sample (65). This is broadly defined as antibiotic tolerance. The MDK assay is used to quantify the spectrum of antibiotic susceptibility within subpopulations based on the duration of the drug exposure required for killing 90-99.99% of the bacterial population. Thus, MDK is a quantitative tolerance assay to measure the antibiotic tolerance of bacterial populations (65). We obtained the MDK_90_, MDK_99_ and MDK_99.99_ of RIF and selected herbal medicinal agents (after exposure for 4-10 days) by using a 96-well plate-based MPN assay. The MPN assay was performed as described (65). The bacterial growth in the 7H9T medium in the 96-well plate was detected by alamar blue assay. The MPN value was either determined by the traditional method as described (65), or by using the extreme limiting dilution (ELDA) statistics (19)(56). In this manner, the MDK_99.99_ dose for RIF was found to be 4 ug/ml/2 days for H37Ra growth, whereas for Ursolic acid MDK_99.99_ dose was 15 ug/ml/4 days.

## 11. Detection of viable but non-culturable (VBNC) MTB by a modified MPN assay

Some dormant bacteria including M. tuberculosis can enter into persistent cells and characterized by not being able to grow in nutrient media even after the removal of stress. These VBNC phenotypes require a much longer time and specific stimuli such as resuscitation factors for their reactivation (66–68). On the other hand, persistent bacteria stochastically arise within growing bacterial culture with or without exposure to stress; they are tolerant to high-dose antibiotics and of growth-arrested phenotype (66). Persisting bacterial population can grow on nutrient media after the removal of the stress, where VBNC phenotype cannot grow on nutrient media even after the removal of stress (66–68). So far, there are no adequate methods to detect VBNC in sputum-negative patients’ sputum, although different assays have been attempted (68–69). We found that the MPN assay described above can be used to determine the VBNC in MTB culture. Briefly, H37Ra were transformed to VBNC by using in vitro hypoxia, as well as macrophage culture method as described (19,56) The VBNC phenotype was confirmed by performing flow cytometry. The VBNCs were then subjected to MPN assay. The resuscitation was achieved in the MPN assay by adding resuscitation factor (RF) to the 7H9 broth and then maintaining the culture for 4-7 days. RF dosage was evaluated using the Alamar Blue (AB) Assay. For MPN assay, the RF-added samples were then serially diluted in the 7H9 broth, and subjected to MBAB assay. The VBNC bacterial solutions were first diluted as follows: 10-1 (1×10^6 bacteria), 10-2, 10-3, 10-4, and 10-5 in a 100 l aliquot. The diluents were mixed into 100 l liquid broth and cultured in 48 well plates under sealed paraffin for 40 days (the paraffin was removed every 10 days and 20 ul of broth was added to maintain the volume). On day 36, 10% Alamar blue was added, and the presence of MTB was identified by visual detection of a change in Alamar blue colour on day 40 (59). AFB staining was used to confirm the presence of MTB in several random wells. The ratio of “resuscitable” to total cell number was obtained in this manner. For the MTB-ELDA method, VBNC suspensions were initially incubated in liquid broth for 7 days in RF to allow VBNC MTB resuscitation. The total number of MTB in the lysate was then determined using AFB staining. Following that, a dilution of 1/10/100/1000 bacteria per 20 ul was made to allow us to undertake the Surface drop method (Miles and Misra method) of agar plating. Thus, tubes containing 5×105, 5×104, 5×103, 5×102, and 5×101 MTB per ml were prepared. 20 ul volume was taken out from each tube and plated as one drop in the Agar plate (Miles and Misra method), resulting in 8 drops on each plate. After 2-3 weeks of culture, positive cultures after each MTB dose (1/10/100/1000/10000) were recorded and the MPN value was calculated by ELDA. The self-renewing MTB per total bacterial count was then computed.

## 12. Bacterial live/dead assay

This is a critical assay for the VBNC estimation. It can be done by BAC light reagents and or EB/FAD staining. For this purpose, bacterial pellets treated with hypoxia or drugs for 4-72 hours are stained with EB and FAD reagents or Bac light reagents, and the vials are covered with aluminium foils for 15 minutes of incubation at 37^0^C. Then, the bacterial samples are subjected to microscopic examination using a Confocal microscope or FACS analysis. The excitation/emission for FAD is done at 480/500 nm and for propidium iodide (red fluorescence dye) at 490/635 nm. The green denotes live, whereas red denotes dead bacteria.

## Discussion

Though tuberculosis has been a major global concern for decades, so far, the worldwide methods to control TB have not been satisfactory. The ineffectiveness of the TB control programs partly contributed to the gradual evolution of the MTB strains (both drug-sensitive and MDR) and their circulation in the identified TB (geographical) hotspots, such as India (2). Recently, there have been reports of misdiagnosis of TB incidences using the standard guidelines (60)(61). Considering the diverse geographical distribution and population of India, and the local issues that hamper implementing, a top-down, centrally controlled TB control programme, there is a need to develop PAR-based approach that can complement our global efforts to eradicate TB.

Taking into consideration all the aspects of this disease pathogenesis (clinical and *in-vivo*/*in-vitro* findings) is vital for implementation of successful treatment plans. Addressing the major shortcomings in the previous TB control methods, such as the lack of public awareness, late detection, and inaccurate diagnosis, our main motives were to evaluate the diagnostic-related logistic issues in remote areas, disease cause evaluation, the role of host-pathogen interaction, patient-specific treatment plans and mass-affordability.

Hence, in our pilot study, we implemented a comprehensive approach comprising all of the steps starting from early screening to routine follow-up post-treatment completion (attached excel sheet), as well as whole-genome sequencing (WGS) to start a database formation for the mapping of circulating strains, and MDR evolution.

We hypothesized that establishing a patient network to understand TB disease spread in a region is essential and should be the first step. Using our previously developed IKS model, a CBPR helped us to effectively trace the TB patients and their contacts. Based on our current findings mentioned in several published, pre-print and unpublished works described in this manuscript, we speculate that the CBPR and IKS-based methodologies may help develop a novel TB control program that can enhance community participation in research including the research on MDR evolution. Thus, there is a strong need to develop CBPR and IKS-based initiatives in rural pockets of India and other developing countries affected by TB. KaviKrishna has been able to implement and sustain the CBPR approach in a non-profit manner for three decades because of the deep connection with the silk weavers and artisans community that started this organization during early part of the 20^th^ century. Importantly, to develop the IKS-based TB control and research program, we were inspired by a historical communication system of the silk industry in Sualkuchi, Assam, India. The communication system was based on a skill-development hub and the establishment of a communication network throughout the Vedic Silk Road (the North-South Silk road or Horse-tea road that stretched from central Asia to Chittagong port in the present Bangladesh; the road went through Bhutan, and Sualkuchi-Hajo cultural complex) to aid in the flow of small-scale silk-weaving projects (15). Such indigenous communication networks are prevalent in many parts of India and Africa, and these networks could be re-activated by our CBPR-IKS methodologies. We noted that one of the key to reactivate and sustain the network is to take a non-profit and non-governmental approach in establishing a health clinic, and diagnostic as well as research laboratory, where the young and energetic students of the community could undergo graduate development programs for their chosen career development. For this purpose, we have set up the CBPR-based clinic, the KaviKrisna Telemedicine care (KTC), a tissue culture hub laboratory, the KaviKrishna lab (KKL) and also a CBPR-based medical humanities program, the center of Indian Knowledge System (K-IKS). So far more than 50 students of the community benefited from the graduate development training program conducted at KaviKrishna since 2010. Next, we have initiated national and international communication networks through collaborative research works with advanced laboratories around the world to serve the local community as well as to move science forward globally. Social network analysis of our activities and network mapping revealed that we were able to develop a bridging-HUB network construct of PAR (15,62), which has made it possible to successfully continue with TB care and research initiatives for the last two decades.

Stigma associated with TB remains a major obstacle in implementing a CBPR-based approach. Even in the 21^st^ century, patients who contract tuberculosis in the rural communities of India are shunned by their society due to prevailing social stigmas and so often these patients go undetected, continuing to spread the disease. However, through a coordinated approach using the temple network, consisting of religious leaders like the Atois, Satradhikars, etc, involvement of student volunteers and KTC team members helped in motivating the patients to come out in open and also trace out their contacts. The regular gatherings of the local people in the Namghars generated awareness of the disease as well as measures to prevent its spread, which also helped us to trace patients and their contacts. Thereafter, we established an efficient registration system for the patients using our KTC database, collected samples, and conducted regular follow-ups. Briefly, after collection, the patient samples were then transferred to KaviKrishna Laboratory (KKL), for molecular biology based diagnostic evaluation and concurrent research on the disease pathogenesis. Resident doctors then analysed the results and gave patients treatment plans accordingly, and encouraged them to continue with the RNTCP-guided treatment plan. At the same time, the biomedical research team at KKL processed the samples for further clinical research to better understand the disease pathogenesis mechanism, host-pathogen interaction, MTB circulating strains and the dormancy-reactivation potential case-by-case. The local students participate in the process through our graduate development program. Local philosophers engage with us in developing experimental design (19). In this manner, the stigma against TB could be minimized. Thus, our PAR serve as a continuous feedback between us and the community leading to the use of IKS to contribute in our knowledge generation of the disease pathogenesis.

These CBPR-based efforts helped us to gain insight on the stem cell basis of TB dormancy and reactivation. Moreover, as per our previous study, we speculated that the dormancy model route utilized by MTB bacteria for disease pathogenesis can play a pivotal role resulting in active TB cases. The ability of viable dormant MTB bacteria to reside in the hypoxic microenvironment of bone marrow MSCs needs to be addressed urgently to prevent dormancy reactivation. The present treatment regime can target the viable MTB present in the macrophages but, not the dormant bacteria. Hence, we evaluated the host-pathogen interaction in this NE cohort by evaluating the BM-MSCs as the site for dormancy and reactivation. These studies through CBPR are helping us not only to understand the basic science but also to understand the epidemiological components of TB dormancy and reactivation. Furthermore, the gradual development program of the CBPR helped us to train a few outstanding students of the community to work on in vitro models to strengthen our clinical findings. Thus, through them, we conducted the *in-vivo* studies to delineate the possible mechanism of dormant MTB reactivation (7). In the near future, we intend to deep dive into these clinical and *in-vivo* findings that can possibly delineate the molecular signalling pathways of dormancy reactivation using stem cell-based studies both at KKL and Thoreau Lab for Global Health at Lowell, MA. Targeting these molecular pathways can eventually eliminate the dormant MTB from their hiding niche and prevent reactivation, thus taking forward our mission for TB eradication.

A key component of our research is the mapping of prevalent strains of MTB circulating in the community by establishing the standard methods as well as the whole genome sequencing. The latest whole genome sequencing analysis has enabled us to surmise the prevalence of the circulating strains in the region and its effect on the patients. The mutation points and their frequency rates also gave us a perspective of the strain viability in the particular area. These findings will now lead our clinicians to curate better treatment plans and recovery regimes for individual patients. The data was further reiterated in elucidating patient-specific disease relapse incidences and re-evaluation of failed treatment regimes. This allowed us to develop generative feedback on the shortcomings of the TB care approach and accordingly streamlining the whole TB control program methodology. The available sequencing data deposited to NCBI data base can be utilized for further in-silico analysis such as advanced epidemiological surveillance analysis and drug developments.

Based on the efficacy of the results obtained by our team (manuscript under preparation), we suggest this overall CBPR based methodology can potentially serve as an effective model for developing novel TB control strategy not only in India but also in the Global south.

In summary, here we have summarized the key methodological aspect of our ongoing CBPR-based screening, diagnosis, treatment monitoring, and research on TB. Here, we also report a pilot study data on WGS and its incorporation in our clinical and research endeavours. Our work suggests the potential role of CBPR-based TB care programs in eradicating this dreaded disease of mankind.

## Data Availability

The excel file of patient follow-up data is available as Supplementary file.

The WGS raw data are available in the NCBI database: Accession no. PRJNA1054003.

## Ethical permission

The clinical study was approved by KaviKrishna Clinical Research and Institutional Ethics Committee. Necessary consents from the patient subjects of Sualkuchi-Hajo area was taken prior to induction for the clinical study. The population represents an extremely homogeneous background. The *in-vivo* and and stem cell research based studies were approved by the Institutional committee for stem cell research (ICSCR), KaviKrishna and Institutional Bio-safety committee (IBSC), KaviKrishna.

## Author Contributions

BD conceptualized and initiated the study. SM, PJS, and LP performed various laboratory experiments with GD’s assistance. LP, RD and SD worked with BD to analyse the philosophical insight of the IKS-based PAR. SM did bioinformatics. AD and BD did the clinical evaluation, patient selection and sample collections with the support of LP, TB, RD, CD and TS. AD with the support of TB, RD, CD and TS did clinical monitoring and care. SM, PJS, AD and LP wrote the manuscript. BD and SD edited the manuscript. All the authors approved the manuscript content.

## Acknowledgment

We thank the members of Department of Stem Cell and Infectious Diseases, KaviKrishna Laboratory, Research Park, Indian Institute of Technology (Guwahati, Assam, India), KaviKrishna Telemedicare Care (Sualkuchi, Assam, India), KaviKrishna Indigenous Knowledge Systems Research Center (Sualkuchi, Assam, India) Department of Stem Cell and Infection, Thoreau Lab for Global Health, University of Massachusetts (Lowell,MA, USA).

